# Impact of Severe COVID-19 on Accelerating Dementia Onset: Clinical and Epidemiological Insights

**DOI:** 10.1101/2025.03.26.25324688

**Authors:** Sasha Mukhija, Max Sunog, Colin Magdamo, Mark W. Albers

**Affiliations:** Neurology Department, Massachusetts General Hospital, Boston, USA; Laboratory of Systems Pharmacology, Harvard Medical School, Boston, USA; Universitätsspital Zürich, Zürich, Switzerland

## Abstract

**Importance:** Severe COVID-19 infection has been associated with neurological complications, but its role in accelerating cognitive decline remains unclear.

**Objective:** To determine whether individuals hospitalized for severe COVID-19 exhibit a higher incidence of new onset cognitive impairment compared to those hospitalized for other conditions.

**Design:** A retrospective study emulating a target trial using Mass General Brigham electronic health records (March 2020-August 2024). The causal effect of COVID-19 hospitalization was estimated via cumulative incidence functions accounting for the competing risk of death.

**Setting:** Multicenter hospital-based study across the Mass General Brigham healthcare system.

**Participants:** A total of 221613 hospitalized patients met the eligibility criteria, including 6454 (2.0%) admitted due to COVID-19 and 215159 (98.0%) for all other conditions. Patients were excluded if they had less than three months of follow-up (due to censoring, cognitive impairment, or death), were younger than 55 years at baseline, or had no prior visit to Mass General Brigham in the year before baseline.

**Main Outcomes and Measures:** The primary outcome was new-onset cognitive impairment, identified via ICD codes and dementia medication prescriptions. The primary analysis estimated the hazard ratio for cognitive impairment with COVID-19 hospitalization relative to other hospitalizations, along with the risk difference at 4.5 years estimated via cumulative incidence functions. Inverse propensity score weighting was used to balance covariates (age, sex, comorbidities, hospitalization period).

**Results:** Among eligible patients (mean [SD] age, 69.55 [9.42] years, 55% female), those hospitalized for COVID-19 were significantly older and had more comorbidities (p < 0.05). COVID-19 hospitalization was associated with a higher risk of developing cognitive impairment (Hazard Ratio: 1.14 [95% CI: 1.02-1.30], P = 0.018). At 4.5 years, the cumulative incidence of cognitive impairment was 12.5% [95% CI: 11.3-13.5] in the COVID-19 group, compared to 11.6% [95% CI: 11.1-12.1] in the non-COVID-19 group.

**Conclusions and Relevance:** Severe COVID-19 infection was associated with an elevated risk of developing clinically recognized cognitive impairment. Future studies are needed to validate findings in other health care settings. Early screening and intervention for cognitive decline may help optimize long-term outcomes for COVID-19 patients.

**Key points:** *Question:* Do individuals hospitalized for severe COVID-19 have a higher incidence of new-onset cognitive impairment compared to those hospitalized for other conditions?

*Findings:* In this retrospective study of 221613 hospitalized patients, including 6454 hospitalized due to COVID-19, a higher incidence of new-onset cognitive impairment was observed among COVID-19 patients.

*Meaning:* Early recognition and diagnosis of dementia in individuals with severe COVID-19 are crucial for optimizing long-term patient outcomes.

## Introduction

COVID-19, caused by the SARS-CoV-2 virus, emerged in late 2019 as a global pandemic with profound and widespread effects on human health. The virus has infected more than 700 million people worldwide^1^ and primarily spreads through respiratory droplets^2^. Infection occurs when non-infected individuals are exposed to these droplets via inhalation or contact with contaminated surfaces^2^. Initially characterized by acute respiratory symptoms, it soon became evident that the virus affects multiple organ systems^3^, leading to a diverse array of complications and even long-term consequences^4^.

Long COVID, also known as PASC (post-acute sequelae of SARS-CoV-2), has affected about 70 million people worldwide^5^l. The World Health Organization defines PASC as a condition where symptoms last at least 3 months after the acute COVID-19 infection and cannot be explained by an alternative diagnosis^6,7^. Symptoms described as “Long COVID” include fatigue, cognitive dysfunction and issues with memory, often referred to as “brain fog”^8^. Long-Covid is more prevalent in people who were hospitalized and suffered from severe Covid-19 infection.^9–11^ As the pandemic progressed, the impact of COVID-19 on the nervous system garnered significant attention^12–14^. A substantial proportion of patients have reported neurological and cognitive symptoms, ranging from mild headaches to severe neuronal degeneration^15^. SARS-CoV-2 is highly pathogenic, capable of infecting various cell types and tissues. However, it remains unclear whether the diverse symptoms result from viral persistence in infected tissues, dysregulated systemic inflammation, or widespread microangiopathy that often leads to microcirculatory thrombi ^4,16^.

In approximately one-third of patients suffering from severe COVID-19, neurological symptoms occur. These symptoms include headaches, paresthesias, strokes, smell loss and neuronal degeneration^4,12,17,18^.

While mounting evidence indicates that SARS-CoV2 is not neurotropic, SARS-CoV2 infection of the respiratory tract has been proposed to affect the CNS through several indirect mechanisms, including angiopathy and persistent neuroinflammation^19^.

SARS-CoV-2 enters the body by attaching to healthy cell membranes via the angiotensin-converting enzyme 2 (ACE2) receptor, which is present in many organ systems^20^. A proposed mechanism for SARS-CoV-2-related olfactory dysfunction involves direct viral targeting of non-neuronal sustentacular support cells in the nasal epithelium that express both the ACE2 receptor and a host cell protease, TMPRSS2 (transmembrane protease serine 2). Once infected, these cells may disrupt the electrophysiological and biochemical homeostasis of nearby olfactory sensory neurons leading to smell distortion. The invasion of SARS-CoV-2 into healthy cells reduces the bioavailability of ACE2 receptors, potentially leading to organ injury through a cytokine storm—an uncontrolled, non-specific inflammatory response—along with increased coagulation, fibrinolysis, and myocardial injury^21,22^. Additionally, conventional MRI scans of individuals with COVID-19 have revealed localized abnormalities in olfactory-eloquent brain regions, suggesting selective susceptibility^4,23^. On the other hand impaired olfaction has been shown to be associated with faster cognitive decline^24^.

The mechanisms underlying memory impairment associated with COVID-19 are not yet fully understood, though several hypotheses have been proposed. Neuroinflammatory markers have been identified in the brains of more than 80% of COVID-19 patients, suggesting that inflammatory processes may contribute to the observed neurological symptoms^25^. It is postulated that viral particles or cytokines may enter the brain by direct transport along the neuronal pathways from the nasal cavity through the cribriform plate and into the olfactory bulb^14,26^. While sudden-onset anosmia or hyposmia (complete or partial loss of smell) has been widely reported as a specific indicator of early infection, the exact mechanisms by which the olfactory system is impaired remain unclear^4^. Interestingly, hyposmia is also an early symptom of Alzheimer’s disease, highlighting a potential overlap in pathophysiologic mechanisms^26^.

Emerging evidence also indicates that a pathway leading to tau hyperphosphorylation, typically associated with Alzheimer’s disease, is activated in COVID-19 patients. This activation is believed to occur through inflammatory signaling by activating the NLRP3 inflammasome and oxidative stress triggered by SARS-CoV-2 infection^27,28^. In general dementia related biomarkers have been found in patients with acute or severe SARS-CoV-2 infection^29–31^. This includes reduced levels of plasma Aβ42, elevated levels of phospho-tau181 and neurofilament light chain – markers associated with Alzheimer’s disease in blood or cerebrospinal fluid^32,33,34^. This further emphasizes the potential long-term neurological impacts of the viral infection of the nasal epithelium^35^. With emerging evidence of increased biomarkers of AD and other neurodegenerative diseases following SARS-CoV2 infection, we tested the hypothesis that SARS-CoV2 infection would increase the risk of being diagnosed with cognitive impairment in subsequent years.

## Methods

### Study population

This study is a multi-center, EHR-based, retrospective target trial emulation (TTE). Patients aged 55 years and older who were hospitalized at Mass General Brigham (MGB) in Boston, USA, between March 1, 2020, and April 29, 2024, were included. After applying exclusion criteria to the total of 1292054 patients hospitalized, 221613 patients remained (Figure 1). The primary outcome was the diagnosis of a new-onset cognitive impairment. For the purposes of this analysis, new onset cognitive impairment includes all forms of clinically diagnosed dementia, as well as milder forms of cognitive impairment, including mild cognitive impairment (MCI) and documented memory loss (Supplementary File 1). This was determined based on a union between International Classification of Diseases (ICD) codes ICD-9 and ICD-10 expertly curated, for memory-related conditions such as memory loss, mild cognitive impairment, Alzheimer’s disease, and related dementias (see Supplementary File 1), as well as the prescription of a dementia medication - galantamine (Razadyne, Razadyne ER), donepezil (Aricept), rivastigmine (Exelon) or memantine (Namenda). Demographic characteristics, comorbidities, and the time periods of hospitalization during different SARS-CoV-2 strain predominance among the included 221613 hospitalized patients at Mass General Brigham were compared. This included 6454 (2.0%) patients admitted due to COVID-19 and 215159 (98%) patients hospitalized for other reasons. Patients hospitalized due to COVID-19 showed significant differences in several characteristics compared to those hospitalized for other reasons (Table 1). Use of data for this study was approved by the institutional review board (IRB) of MGB (Protocol 2023P000604).

**Figure 1.**
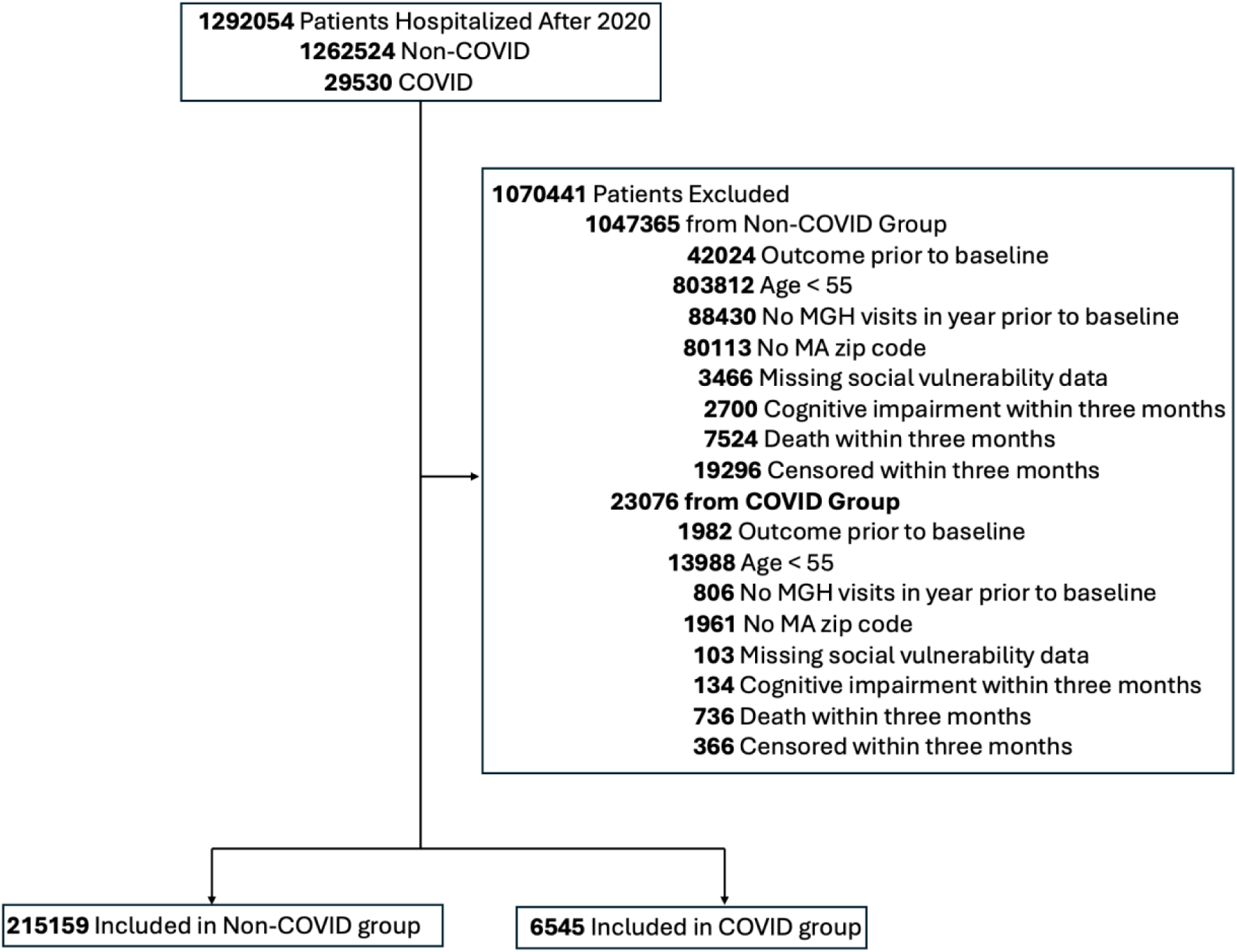
Consortium Diagram with Inclusion and Exclusion Criteria.

**Table 1.** Demographics, Comorbidities, and COVID-19 Strain Periods Among COVID-19 Hospitalized Patients vs. Non-COVID Patients Hospitalized for All Other Reasons.

| Covariates | Hospitalized for all other reasons (n= 215159) | Hospitalized due to COVID-19 (n=6454) | p |
| --- | --- | --- | --- |
| No. (%) female | 118080 (55%) | 3335 (52%) | <0.001 |
| Age, mean (SD), years | 69.53 (9.40) | 70.29 (9.98) | <0.001 |
| No. (%) with hypertension | 122406 (57%) | 4003 (62%) | <0.001 |
| No. (%) with stroke | 9590 (4%) | 280 (4%) | 0.649 |
| No. (%) with COPD | 15168 (7%) | 577 (9%) | <0.001 |
| No. (%) with obesity | 80181 (37%) | 2673 (41%) | <0.001 |
| No. (%) with overweight | 148371 (69%) | 4474 (69%) | 0.535 |
| No. (%) with cardiovascular disease | 45710 (21%) | 1543 (24%) | <0.001 |
| No. (%) with diabetes | 38106 (18%) | 1629 (25%) | <0.001 |
| No. (%) with graduate education | 23469 (11%) | 535 (8%) | <0.001 |
| No. (%) college education | 90624 (42%) | 2273 (35%) | <0.001 |
| No. (%) with missing education | 40023 (19%) | 1182 (18%) | 0.559 |
| No. (%) during Time Period of Strain-Alpha (October 2020-June 2021) | 43371 (20%) | 1588 (25%) | <0.001 |
| No. (%) during Time Period of Strain-Delta (July 2021-November 2021) | 23559 (11%) | 330 (5%) | <0.001 |
| No. (%) during Time Period of Strain-Omicron (December 2021-February 2022) | 10146 (5%) | 712 (11%) | <0.001 |
| No. (%) during Time Period of Strain-Post Omicron (March 2023-end of analysis August 2024) | 85667 (40%) | 2682 (42%) | <0.001 |
| No. (%) during Time Period of Strain- Pre Alpha (March 2020-September 2020) | 52416 (24%) | 1142 (18%) | <0.001 |

### Target Trial Emulation

Causal effects of interventions are typically tested through randomized controlled trials (RCTs), but RCTs are not always feasible. For example, assigning participants to a COVID-19 infection would be unethical, making an RCT impossible. Instead, the causal effect of COVID-19 hospitalization versus hospitalization for other reasons can be examined using the TTE framework, which retrospectively mimics an RCT using electronic health records.

In a TTE, eligible patients are identified based on predefined criteria in the EHRs. Their records determine the baseline date and treatment assignment (e.g., COVID-19 hospitalization), while follow-up encounters establish outcomes or censoring dates. Since treatment arms in a TTE are not randomized, inverse propensity of treatment weighting (IPTW) is applied to reduce confounding. IPTW ensures that observed differences are attributable to the intervention rather than preexisting factors, provided all relevant confounders are measured and no patient is entirely ineligible for either treatment arm.^36,37^

### Statistical assessment

Data for this study were obtained from the Enterprise Data Warehouse (EDW) at Mass General Brigham using Microsoft Structured Query Language (SQL) Server Management Studio. Secondary analyses were conducted using RStudio (Version 4.3).

To account for potential confounding factors and ascertain the causal effect of interest, IPTW was used to balance the group hospitalized due to COVID-19 with the group hospitalized for other reasons across several covariates, including sex, age, comorbidities (e.g., hypertension, diabetes), and period of hospitalization (Pre-Alpha, Alpha, Delta, Omicron, and Post-Omicron Covid Strain). The efficacy of the propensity score weighting is demonstrated through the successful balancing of covariates between the groups. The primary outcome of interest was the diagnosis of new onset cognitive impairment, and the statistical significance of differences between groups was assessed using hazard ratios (HRs) with 95% CIs derived from Cox proportional hazards models^38^.

The competing risk of death was accounted for in the cumulative incidence functions, which estimated the probability of new-onset cognitive impairment and of death before cognitive impairment over a 4.5-year period^39^.

## Results

Out of the 221613 patients included in the study, 6454 (2%) were hospitalized due to COVID-19. Patients hospitalized due to COVID-19 have a significantly higher risk of developing new onset cognitive impairment compared to those hospitalized for other reasons. The hazard ratio (HR) for cognitive impairment among hospitalized COVID-19 patients was 1.14 (95% CI: 1.02– 1.28; *p* = 0.017) on average over 4.5 years of follow up (Figure 2).

**Figure 2.**
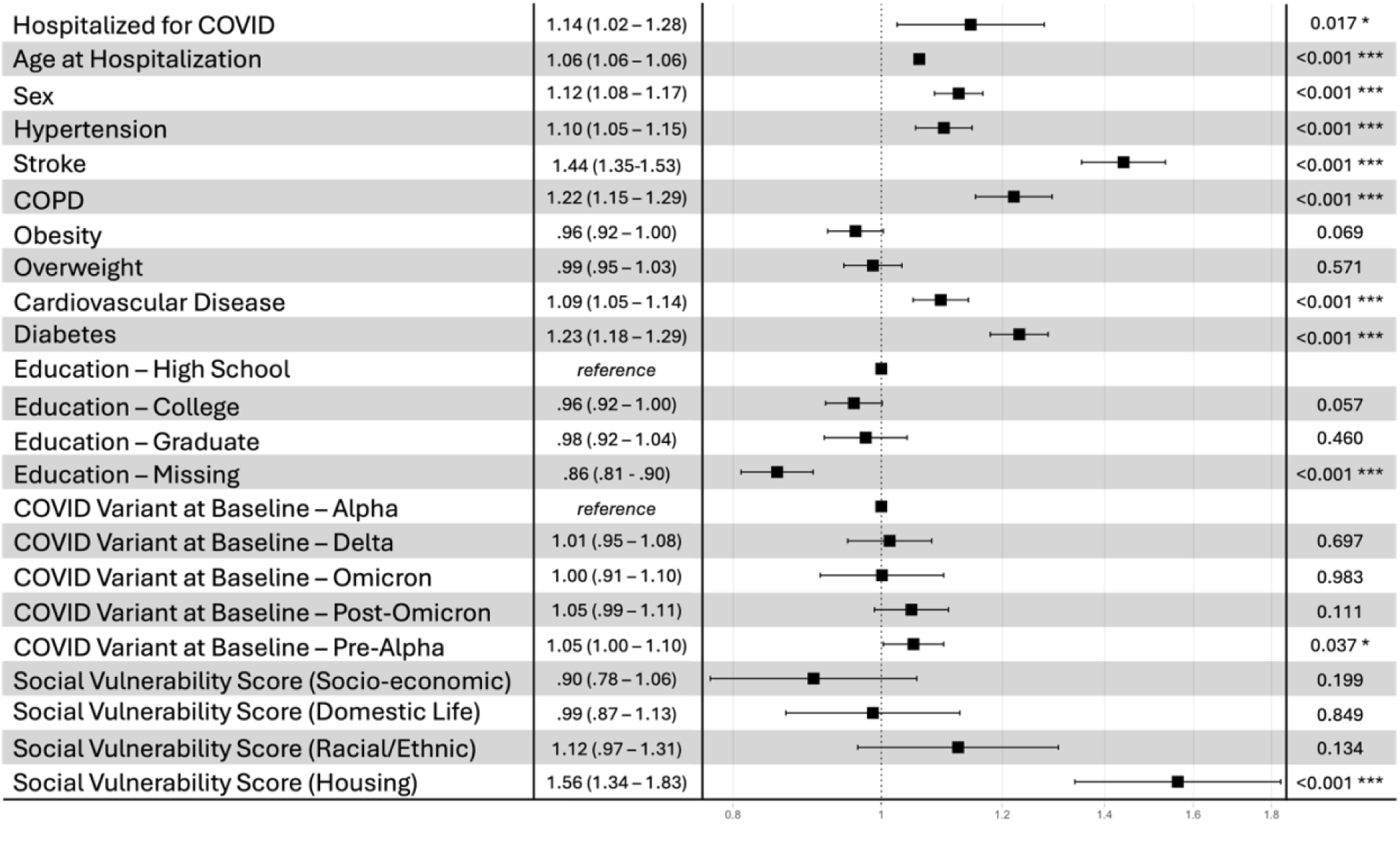
Hazard Ratios for Dementia Risk in COVID-19 vs. Non-COVID-19 Hospitalized Patients Across Covariates.

Increasing age conferred a significant increase in risk of cognitive impairment risk (HR 1.06 per year, 95% CI 1.057–1.061, p < 0.001), and female sex was also associated with elevated risk (HR 1.12, 95% CI 1.08–1.17, p < 0.001).

Among comorbidities, COPD (chronic obstructive pulmonary disease) (HR 1.22, 95% CI 1.15– 1.29, p < 0.001), diabetes (HR 1.23, 95% CI 1.18–1.29, p < 0.001), cardiovascular disease (HR 1.09, 95% CI 1.05-1.14, p>0.001), hypertension (HR 1.10, 95% CI 1.05–1.15, p < 0.001), and history of stroke (HR 1.44, 95% CI 1.35–1.53, p < 0.001) were each significantly associated with greater risk of cognitive impairment risk. In contrast, being merely overweight (HR 0.99, 95% CI 0.95–1.03, p = 0.571) and being obese (HR 0.96, 95% CI 0.92–1.00, p = 0.069) were not statistically significant predictors.

Among those with recorded educational status, the level of attainment had no significant effects. Compared to high school education, a college-level education correlated with a decreased hazard of cognitive impairment (HR 0.96, 95% CI 0.92–1.00, p =0.057), whereas a graduate-level education was marginally protective (HR 0.98, 95% CI 0.92–1.04, p = 0.460). Missing data on education showed a significant effect (HR 0.86, 95% CI 0.81–0.90, p <0.001).

Finally, relative to the reference period which is the time during the Alpha-strain of SARS-CoV- 2, the pre-Alpha era was associated with a significantly higher increased risk for cognitive impairment (HR 1.05, 95% CI 1.00–1.10, p = 0.037), whereas the Delta-period estimate (HR 1.01, 95% CI 0.95–1.08, p = 0.697) and subsequent viral waves (Omicron, Post-Omicron) did not show consistent or significant deviations from the reference risk (CIs crossing or approaching unity).

In summary, COVID-19 hospitalization was associated with a significant elevation in the hazard of new onset cognitive impairment, over and above known risk factors such as older age, comorbid cardiopulmonary conditions, and diabetes.

The propensity score-weighted analysis demonstrated effective balancing of covariates.

After adjusting for the competing risk of death, the cumulative probability of being diagnosed with cognitive impairment over a 4.5-year follow-up period was higher among patients hospitalized due to COVID-19 compared to those hospitalized for other reasons. Specifically, the probability of developing cognitive impairment was 12.5% (95% CI: 11.3%–13.5%) in the COVID-19 cohort, compared to 11.6% (95% CI: 11.1%–12.1%) in the cohort hospitalized for other conditions (Figure 3). The COVID-19 cohort exhibited a cumulative incidence of death of 16.2% (95% CI: 15.0%–17.6%) over the follow-up period, compared to 16.3% (95% CI: 15.5%– 17.1%) in the non-COVID hospitalized cohort.

**Figure 3.**
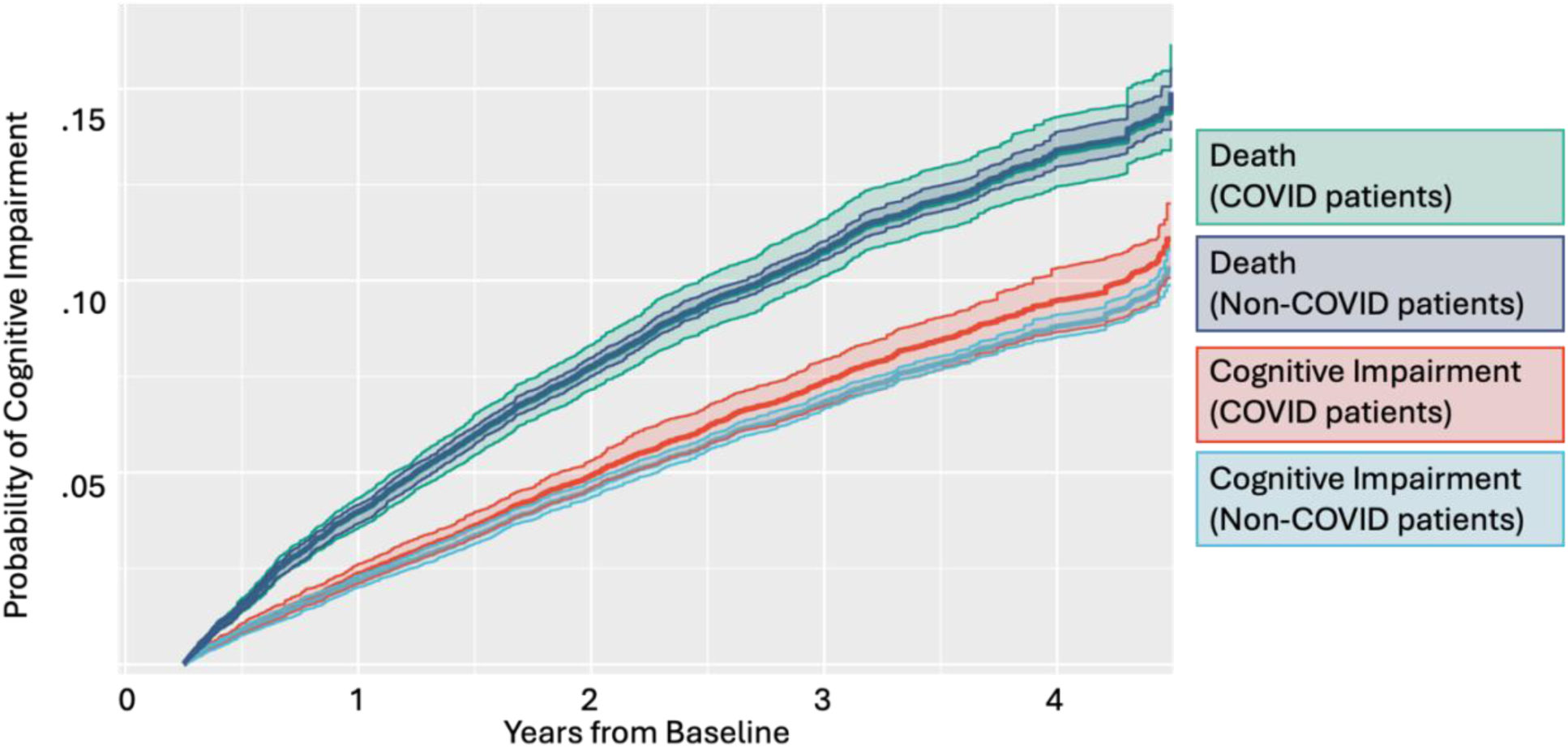
Cumulative Probability of Incident Cognitive Impairment and Death Over a 4.5-Year Period for COVID-19 vs. Non-COVID-19 Hospitalized Patients.

## Discussion

Using causal inference approaches, we tested the hypothesis that patients hospitalized due to COVID-19 were more likely to develop a cognitive impairment relative to patients who were hospitalized concurrently for other reasons. After balancing the two treatment arms using IPTW and controlling for the competing risk of death, we found that patients with severe COVID had an elevated risk of being diagnosed with cognitive impairment 3 months or later after baseline, relative to patients concurrently hospitalized for other reasons. The divergence between the cumulative incidence curves for new-onset cognitive impairment between the two groups becomes particularly evident after year 2, suggesting that the long-term neurological impact of COVID-19 hospitalization may manifest progressively over time.

Importantly, these results were derived using a competing risk model, which accounts for the likelihood of death as a competing event. This adjustment ensures that the observed differences in risk for cognitive impairment are not confounded by variations in mortality rates between the groups.

When stratifying by cause of hospitalization, individuals hospitalized due to COVID-19 not only had an elevated risk of cognitive impairment but also a greater probability of death.

The competing risk framework highlights that, despite the increased mortality in the COVID-19 group, the risk for cognitive impairment remains significantly elevated, indicating that the observed cognitive decline is not attributable to survival bias. The greater risk of new onset cognitive impairment among those hospitalized with COVID-19, despite the competing risk of death, underscores the long-term neurological consequences of viral infection with severe symptoms. A UK based study of hospitalized COVID-19 patients has identified the most common neurologic conditions as being anosmia, stroke, delirium, brain inflammation, encephalopathy, primary psychiatric syndromes and peripheral nerve syndromes^37^. Many of these neurologic sequelae are also risk factors for age-related neurodegenerative diseases.

These findings support the hypothesis that mechanisms such as neuroinflammation, cerebrovascular complications, and direct viral effects on the brain contribute to cognitive decline in COVID-19 patients. This underscores the critical need for long-term follow-up care and cognitive monitoring for COVID-19 survivors.

Additionally, individuals with pre-existing comorbidities -- such as hypertension and diabetes -- and older adults are at heightened risk, suggesting that these populations may require more intensive surveillance and early intervention. Other studies have also shown that plasma biomarker changes (Aβ42, and phospho-tau181) were larger in patients that were hospitalized due to their COVID-19 infection or had previously diagnosed hypertension^34^. It is postulated that these conditions might involve pre-existing ACE-2 deficiencies predating SARS-CoV-2 infection. As a result, the risk of a more severe infection is increased due to a further reduced bioavailability of ACE2 receptors^40–42^.

While evidence for increased dementia rates following severe SARS-CoV-2 infection has previously been reported ^28,30,43^, our study provides novel contributions in both scale and methodology. By leveraging a large, longitudinal cohort with up to 4.5 years of follow-up and using non-COVID hospitalized controls, we were able to isolate the impact of severe COVID-19 beyond hospitalization alone. Advanced causal inference methods, including inverse propensity score weighting and competing risk modeling, allowed us to address confounding and the competing risk of death.

The findings contribute to a growing body of evidence indicating that COVID-19 may have long-lasting effects on brain health, especially in vulnerable groups. This is particularly concerning given the widespread prevalence and enduring impact of the virus, highlighting the necessity for healthcare systems to adapt and prepare for potential long-term increases in dementia cases.

Moreover, the biological mechanisms linking SARS-CoV-2 infection to cognitive decline merit further investigation. Neuroinflammatory markers identified in COVID-19 patients suggest that inflammatory processes play a crucial role. Studies have shown that COVID-19 could potentially exacerbate neuroinflammation and contribute to neurodegeneration, highlighting the fact that peripheral infection and its subsequent immune response could be responsible for the neuropathology^34,44^.

The observed association between severe COVID-19 and subsequent cognitive impairment underscores the need for continued research to elucidate the underlying pathways. Understanding these connections is essential for developing targeted therapeutic strategies to mitigate the long-term cognitive impacts of COVID-19.

## Limitations

The study’s retrospective design may introduce limitations inherent in the use of EHRs, caused by incomplete data and inaccuracies in coding, which were exacerbated by the disruption caused by the pandemic. To specifically investigate the effect of severe COVID-19, patients were only assigned to the treatment arm if a confirmed COVID-19 infection was listed as a reason for hospitalization. However, patients in the control arm may have had Covid-19 infection that did not lead to hospitalization prior to baseline; mild, untested, or unrecorded COVID-19 infection during their hospitalization; or have contracted Covid-19 in the hospital after admission. While inverse propensity score weighting was employed to balance observed covariates, residual confounding due to unmeasured factors, such as genetic predispositions or lifestyle factors, cannot be entirely excluded. The cohort is predominantly composed of patients from a single healthcare system (Mass General Brigham), which may limit the generalizability of the findings to other populations or healthcare settings. This study may have measurement errors in its primary outcome. Dementia is often underdiagnosed and under-recorded^45^. Patients and families may not report symptoms, and physicians may not routinely screen for cognitive health. Even when symptoms are noted, diagnostic codes or prescriptions used as proxies may be missing or appear late. In some cases, dementia may also be overcoded^46^.

## Conclusion

In conclusion, our findings suggest that severe COVID-19 may accelerate the onset of cognitive impairment after accounting for the competing risk of death. Early clinical recognition and diagnosis of dementia in this population are critical for improving long-term patient outcomes. Screening tools that assess the neural processing of smells in patients who had COVID-19 could serve as an important method for monitoring cognitive function and early neurological changes^47^. At home preclinical cognitive tests, such as BRANCH (Boston Remote Assessment for Neurocognitive Health), may be sensitive to detect subtle declines^48^.

The observed increase in dementia risk, although modest, carries significant clinical implications given the substantial number of COVID-19 hospitalizations globally. It emphasizes the necessity for long-term follow-up care and cognitive monitoring for COVID-19 survivors, especially those at heightened risk due to pre-existing health conditions. Establishing biomarkers for predicting subsequent cognitive decline after SARS-CoV2- infection might be a promising strategy^28^.

Further research is essential to replicate these results in other health care systems and understand the relationship of vaccination to mitigate the risk of dementia. Elucidating the specific mechanisms involved in neurodegenerative disease progression could guide the development of targeted interventions aimed at preventing or mitigating dementia and other long-term neurological diseases. This could improve the quality of life for COVID-19 survivors.

## Data Availability

Researchers can obtain an anonymized version of the study dataset from the authors upon request and completion of the MGB Health data use agreement for the use of EDW data. This agreement ensures the privacy of MGB patients and compliance with US regulatory standards and has been approved by the MGB IRB. In addition, we provide a summary about patient data security and privacy. Patients who visit Mass General Brigham receive a HIPAA notice that states that their identifiable data may be used for research with proper Institutional Review Board approval. The patient has an opportunity to object to this usage of their data by seeking care outside of Mass General Brigham. The IRB classifies aggregate queries of online patient registries that are populated with appropriately obfuscated, de-identified/encrypted data, performed by authorized staff as a category of research that is exempt from IRB review. In addition, HIPAA privacy rules do not apply to de-identified information. Therefore, the Mass General Brigham Research Council determined that faculty, and those overseen directly by faculty, are approved for access to the Query Tool. However, with respect to the Detailed Data Wizard, the IRB must review and approve the release of identified and de-identified medical record data to researchers. In an effort to secure patient privacy and prevent a security breach, all patient identifiers are encrypted throughout the database.

## Conflict of Interest

All authors declare no competing interests or conflicts.

## Author Contribution

Max Sunog and Colin Magdamo had full access to all of the data in the study and take responsibility for the integrity of the data and the accuracy of the data analysis. Sasha Mukhija, Max Sunog, Colin Magdamo and Mark W. Albers designed the study, analyzed data and edited/wrote the paper.

## Funding/Support

S.M. was supported by a Fulbright Fellowship. This study was funded by R01 AG058063 and U01DC019579 (awarded to M.W.A.)

## Role of the Funder/Sponsor

The funders had no role in the design and conduct of the study; collection, management, analysis, and interpretation of the data; preparation, review, or approval of the manuscript; and decision to submit the manuscript for publication.

## Data sharing statement

See Supplement File 2

## Supplementary File 1

| Code | Name |
| --- | --- |
| F03.91 | Unspecified dementia with behavioral disturbance |
| F02.81 | Dementia in other diseases classified elsewhere with behavioral disturbance |
| d290.43.1 | Vascular dementia, With depressed mood |
| d290.42.1 | Vascular dementia With delusions |
| F01.51 | Vascular dementia with behavioral disturbance |
| 290.20 | Senile dementia with delusional features |
| 290.3 | Senile dementia with delirium |
| 290.43 | Arteriosclerotic dementia with depressive features |
| 290.41 | Arteriosclerotic dementia with delirium |
| 290.21 | Senile dementia with depressive features |
| 290.12 | Presenile dementia with delusional features |
| 290.13 | Presenile dementia with depressive features |
| 290.42 | Arteriosclerotic dementia with delusional features |
| 290.2 | Senile dementia with delusional or depressive features |
| 294.21 | Dementia, unspecified, with behavioral disturbance |
| 294.11 | Dementia in conditions classified elsewhere with behavioral disturbance |
| F03.9 | Unspecified dementia |
| 331.1 | Pick's disease |
| d290.0.1 | Senile Dementia Uncomp |
| F02.80 | Dementia in other diseases classified elsewhere without behavioral disturbance |
| F03.90 | Unspecified dementia without behavioral disturbance |
| G31.83 | Dementia with lewy bodies |
| F01.50 | Vascular dementia without behavioral disturbance |
| LPA99 | Dementia-LMR 99 |
| LPA867 | Alzheimer's disease-LMR 867 |
| ODA:YHAL6 | Dementia-Oncall |
| d290.40.1 | Vascular dementia, uncomplicated |
| LPA1009 | Alzheimers disease-LMR 1009 |
| G31.09 | Other frontotemporal dementia |
| G31.01 | Pick's Disease |
| ODA:WHMT3 | Alzheimer's disease-Oncall |
| d331.0.3 | Alzheimer's disease |
| d294.10.1 | Dementia Conditions Classified Elsewhere w/o Behavioral Disturbances |
| d290.10.1 | Presenile dementia |
| F03 | Unspecified dementia |
| 331.19 | Other frontotemporal dementia |
| 331.11 | Pick's disease |
| 331.82 | Dementia with Lewy bodies |
| G30.8 | Other alzheimer's disease |
| G30.9 | Alzheimer's disease, unspecified |
| G30.0 | Alzheimer's disease with early onset |
| G30.1 | Alzheimer's disease with late onset |
| 290.0 | Senile dementia, uncomplicated |
| 290.40 | Arteriosclerotic dementia, uncomplicated |
| 290.10 | Presenile dementia, uncomplicated |
| 290.11 | Presenile dementia with delirium |
| 290 | Senile and presenile organic psychotic conditions |
| 290.4 | Arteriosclerotic dementia |
| 290.1 | Presenile dementia |
| 294.10 | Dementia in conditions classified elsewhere without behavioral disturbance |
| 294.20 | Dementia, unspecified, without behavioral disturbance |
| 294.1 | Dementia in conditions classified elsewhere |
| 331.0 | Alzheimer's disease |
| 331.00 | Alzheimer's disease |
| F02 | Dementia in other diseases classified elsewhere |
| 294.2 | Dementia, unspecified |
| F01 | Vascular dementia |
| G31.1 | Senile degeneration of brain, not elsewhere classified |
| G31.9 | Degenerative disease of nervous system, unspecified |
| 331.2 | Senile degeneration of brain |
| 331.8 | Other Cerebral Degeneration |
| 331.9 | Cerebral Degeneration, unspecified |
| G31.81 | Alpers disease |
| G31.85 | Corticobasal degeneration |
| G31.89 | Other specified degenerative diseases of nervous system |
| 331.89 | Other Cerebral Degeneration |
| 331 | Other Cerebral Degenerations |
| 290.9 | Unspecified senile psychotic condition |
| 290.8 | Other specified senile psychotic conditions |
| 294.9 | Unspecified organic brain syndrome (chronic) |
| 294.8 | Other specified organic brain syndromes (chronic) |
| 294 | Other organic psychotic conditions (chronic) |
| 438.0 | Cognitive deficits as late effect of cerebrovascular disease |
| G31.84 | Mild cognitive impairment, so stated |
| ODA:WLEN6 | Cognitive impairment-Oncall |
| 331.83 | Mild cognitive impairment, so stated |
| F04 | Amnestic disorder due to known physiological condition |
| 780.93 | Memory loss |
| R41.2 | Retrograde amnesia |
| R41.3 | Other amnesia |
| I69.211 | Memory deficit following other nontraumatic intracranial hemorrhage |
| LPA730 | Memory loss-LMR 730 |
| LPA1404 | Memory loss-LMR 1404 |
| ODA:WLAG8 | Memory loss-Oncall |
| I69.311 | Memory deficit following cerebral infarction |
| I69.811 | Memory deficit following other cerebrovascular disease |
| I69.911 | Memory deficit following unspecified cerebrovascular disease |
| 294.0 | Amnestic syndrome |
| R41.81 | Age-related cognitive decline |
| 797 | Senility without mention of psychosis |
| 797.0 | Senility without mention of psychosis |
| G30 | Alzheimer's disease |
| G31.0 | Frontotemporal dementia |
| G31.8 | Other specified degenerative diseases of nervous system |
| F01.5 | Vascular dementia without behavioral disturbance |
| F02.8 | Dementia in other diseases classified elsewhere with behavioral disturbance |
| F01.A | Vascular Dementia, mild |
| F01.B | Vascular Dementia, moderate |
| F01.C | Vascular Dementia, severe |
| F02.A | Dementia in other diseases classified elsewhere, mild |
| F02.B | Dementia in other diseases classified elsewhere, moderate |
| F02.C | Dementia in other diseases classified elsewhere, severe |
| F03.A | Unspecified dementia, mild |
| F03.B | Unspecified dementia, moderate |
| F03.C | Unspecified dementia, severe |

## Notes

### Competing Interest Statement

The authors have declared no competing interest.

### Author Declarations

Use of data for this study was approved by the Institutional Review Board (IRB) of Mass General Brigham (Protocol 2023P000604).

### Summary of Updates

Corrected the spelling of the last name of the first author. No change to the manuscript.

